# T cell anergy in COVID-19 reflects virus persistence and poor outcomes

**DOI:** 10.1101/2020.09.21.20198671

**Authors:** Kerstin Renner, Christine Müller, Charlotte Tiefenböck, Jan-Niklas Salewski, Frederike Winter, Simone Buchtler, Maximilian V. Malfertheiner, Matthias Lubnow, Dirk Lunz, Bernhard Graf, Florian Hitzenbichler, Frank Hanses, Hendrik Poeck, Marina Kreutz, Evelyn Orsó, Ralph Burkhardt, Tanja Niedermair, Christoph Brochhausen, André Gessner, Bernd Salzberger, Matthias Mack

## Abstract

Coronavirus disease 2019 (COVID-19) can lead to severe pneumonia and hyperinflammation. So far, insufficient or excessive T cell responses were described in patients. We applied novel approaches to analyze T cell reactivity and showed that T anergy is already present in non-ventilated COVID-19 patients, very pronounced in ventilated patients, strongly associated with virus persistence and reversible with clinical recovery. T cell activation was measured by downstream effects on responder cells like basophils, plasmacytoid dendritic cells, monocytes and neutrophils in whole blood and proved to be much more meaningful than classical readouts with PBMCs. Monocytes responded stronger in males than females and IL-2 partially reversed T cell anergy. Downstream markers of T cell anergy were also found in fresh blood samples of critically ill patients with severe T cell anergy. Based on our data we were able to develop a score to predict fatal outcomes and to identify patients that may benefit from strategies to overcome T cell anergy.

## Introduction

SARS-CoV-2, the etiologic agent of coronavirus disease 19 (COVID-19), was discovered in Wuhan (China) in December 2019 and became pandemic within a couple of months^1^. Most patients with SARS-CoV-2 infection are asymptomatic or show only mild symptoms, while some patients develop bilateral interstitial pneumonia and require oxygen support by nasal cannula or mechanical ventilation on an intensive care unit (ICU)^2^.

Several risk factors for COVID-19 associated death have been identified including age, gender, adipositas, pulmonary diseases and diabetes^3-6^. Acute respiratory distress syndrome (ARDS) and fatal outcomes are also associated with hyperinflammation and microthrombosis^7^. Activated monocytes / macrophages contribute to death in a mouse model of SARS-CoV infection^8^ and also seem to play an important role for hyperinflammation in human SARS-CoV-2 infection^9,10^. Monocytes are activated by a wide range of signals, including pathogen-derived signals (e.g. nucleic acids) and T cell-derived signals (e.g. cytokines)^11^. The innate immune system seems to be insufficient to rapidly clear SARS-CoV-2 infection by itself, as SARS-CoV-2-specific T and B cell responses develop in most patients^12-14^. Virus-specific T cell responses and neutralizing antibodies, typically generated in a T cell dependent manner, are considered essential for viral clearance^15^. Potential benefits of plasma obtained from convalescent COVID-19 patients also points into this direction^16,17^. On the other hand, hyperinflammation in COVID-19 patients might be caused by excessive T cell responses that activate monocytes and neutrophils. Beneficial effects of dexamethason indicate that hyperinflammation contributes to fatal outcomes^18,19^. However, dexamethason does not only interfere with T cell activation and could also exert therapeutic effects by blocking excessive innate immune responses against viral components.

Current data on T cell reactivity in COVID-19 are conflicting and indicate that patients with severe COVID-19 can have either insufficient^20,21^ or excessive^22-24^ T cell responses, as recently summarized^25^. T cell responses were measured by intracellular cytokine staining of PBMCs, cultured with phorbolesters and ionomycin, by quantification of cytokines after polyclonal stimulation or by expression of surface markers on T cells. IFN-gamma expression for example was found to be unchanged^26^, increased^23,24^ or decreased^20,21^.

We applied novel approaches to analyze T cell reactivity in COVID-19 patients and controls. T cell activation was measured by downstream effects on responder cells like basophils, pDCs, monocytes and neutrophils in whole blood. These assays turned out to be much more sensitive and consistent than classical readouts with PBMCs. We found that impaired T cell reactivity is already evident in mild disease and strongly associated with disease severity and prolonged viral replication. Gender-specific differences in T cell-induced downstream effects on monocytes were observed. Finally we developed a score to predict fatal outcome and identify patients that may benefit from strategies to overcome T cell anergy.

## Results

### Clinical characterization of COVID-19 patients

We analyzed 55 patients with COVID-19 infection admitted to the University Hospital Regensburg from March to July 2020. SARS-CoV-2-infection was diagnosed by RT-PCR from oropharyngeal swabs. Patients were stratified into “non-ventilated” and “ventilated” patients according to the need of mechanical ventilation. Ventilated patients were further sub-grouped into “survived”, if they could be discharged from the ICU, or into “dead” if they died on the on the ICU. In addition, 42 healthy controls were included in the study. Clinical characteristics of patients and controls are shown in Table. 1. In most patients several consecutive blood samples were available resulting in a total of 188 samples. 14 patients were first sampled on mechanical ventilation and also after weaning from ventilation. Samples of these patients were assigned into the subgroup “ventilated” or “non-ventilated” according to the ventilation status at the time of sampling (Supplementary Tab. 1).

**Tab. 1.**
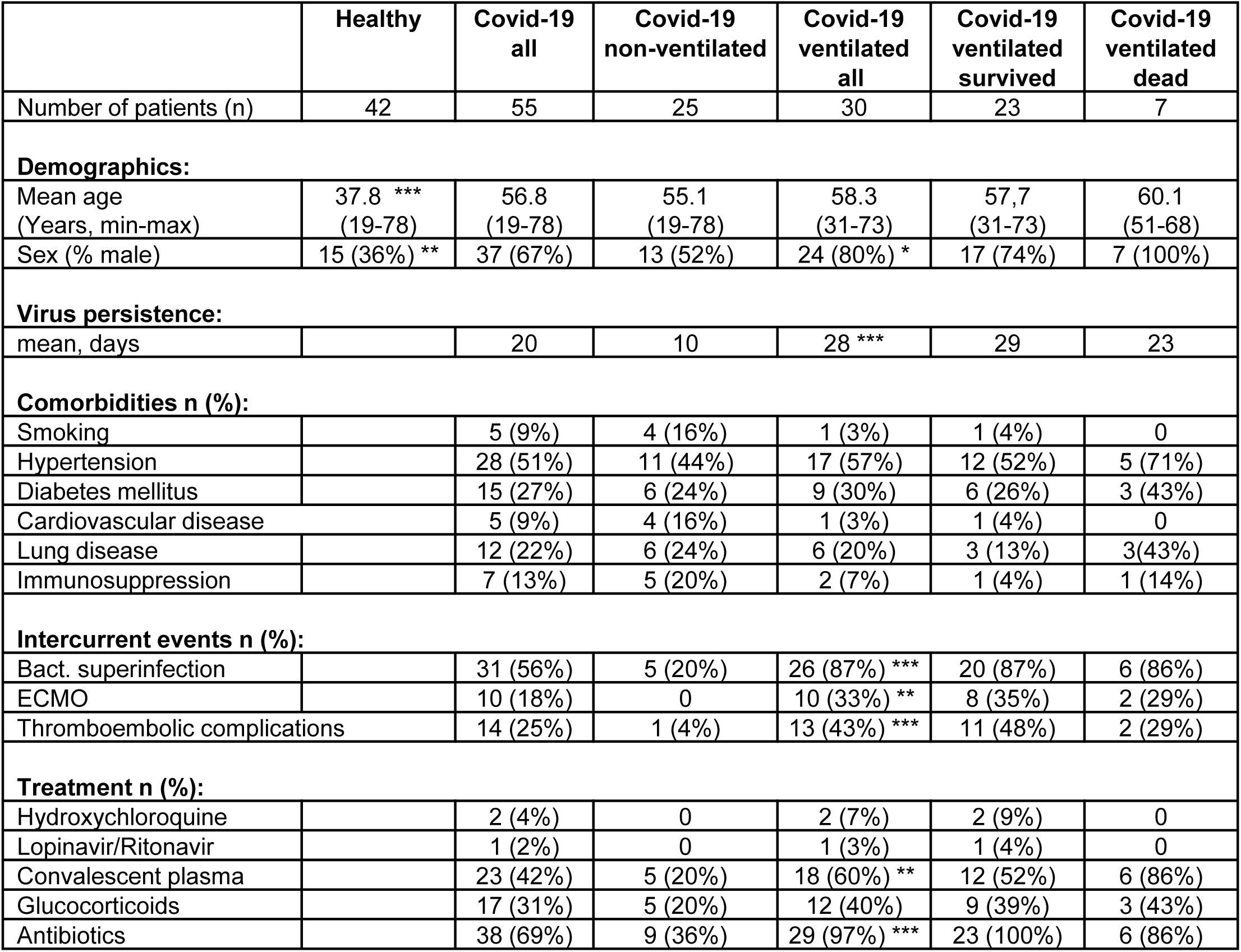
Demographics and clinical information from study patients and healthy controls. COVID-19 patients were subgrouped into non-ventilated and ventilated patients. Non-ventilated patients were hospitalized on a normal ward and received oxygen by nasal cannula as needed. Ventilated patients were treated on an ICU by mechanical ventilation. Ventilated patients were further subgrouped into patients that were discharged from the ICU (“Survived”) and patients that died on the ICU (“Dead”). Virus persistence was calculated form the day of first clinical symptoms to the day of the last positive virus PCR. 2-tailed unpaired t-test was used to calculate statistical differences between non-ventilated and ventilated patients as well as between “Survived” and “Dead” patients. (^*^ p<0.05, ^**^ p<0.01, ^***^ p<0.001).

|  | Healthy | Covid-19<br>all | Covid-19<br>non-ventilated | Covid-19<br>ventilated<br>all | Covid-19<br>ventilated<br>survived | Covid-19<br>ventilated<br>dead |
| --- | --- | --- | --- | --- | --- | --- |
| Number of patients (n) | 42 | 55 | 25 | 30 | 23 | 7 |
| **Demographics:** |  |  |  |  |  |  |
| Mean age<br>(Years, min-max) | 37.8 ***<br>(19-78) | 56.8<br>(19-78) | 55.1<br>(19-78) | 58.3<br>(31-73) | 57,7<br>(31-73) | 60.1<br>(51-68) |
| Sex (% male) | 15 (36%) ** | 37 (67%) | 13 (52%) | 24 (80%) * | 17 (74%) | 7 (100%) |
| **Virus persistence:** |  |  |  |  |  |  |
| mean, days |  | 20 | 10 | 28 *** | 29 | 23 |
| **Comorbidities n (%):** |  |  |  |  |  |  |
| Smoking |  | 5 (9%) | 4 (16%) | 1 (3%) | 1 (4%) | 0 |
| Hypertension |  | 28 (51%) | 11 (44%) | 17 (57%) | 12 (52%) | 5 (71%) |
| Diabetes mellitus |  | 15 (27%) | 6 (24%) | 9 (30%) | 6 (26%) | 3 (43%) |
| Cardiovascular disease |  | 5 (9%) | 4 (16%) | 1 (3%) | 1 (4%) | 0 |
| Lung disease |  | 12 (22%) | 6 (24%) | 6 (20%) | 3 (13%) | 3(43%) |
| Immunosuppression |  | 7 (13%) | 5 (20%) | 2 (7%) | 1 (4%) | 1 (14%) |
| **Intercurrent events n (%):** |  |  |  |  |  |  |
| Bact. superinfection |  | 31 (56%) | 5 (20%) | 26 (87%) *** | 20 (87%) | 6 (86%) |
| ECMO |  | 10 (18%) | 0 | 10 (33%) ** | 8 (35%) | 2 (29%) |
| Thromboembolic complications |  | 14 (25%) | 1 (4%) | 13 (43%) *** | 11 (48%) | 2 (29%) |
| **Treatment n (%):** |  |  |  |  |  |  |
| Hydroxychloroquine |  | 2 (4%) | 0 | 2 (7%) | 2 (9%) | 0 |
| Lopinavir/Ritonavir |  | 1 (2%) | 0 | 1 (3%) | 1 (4%) | 0 |
| Convalescent plasma |  | 23 (42%) | 5 (20%) | 18 (60%) ** | 12 (52%) | 6 (86%) |
| Glucocorticoids |  | 17 (31%) | 5 (20%) | 12 (40%) | 9 (39%) | 3 (43%) |
| Antibiotics |  | 38 (69%) | 9 (36%) | 29 (97%) *** | 23 (100%) | 6 (86%) |

**Tab. 2.** Predictive score for death in ventilated COVID-19 patients. Three parameters were used individually or in combination to predict fatal outcome in ventilated COVID-19 patients. Absolute basophil counts in fresh peripheral blood. Basophil counts < 25 / µl defined “Low Baso count”. Upregulation of CD123 on CD14+ monocytes and CD11b on neutrophils defined as the ratio of surface marker expression with anti-CD3 and surface marker expression without anti-CD3. Upregulation of < 130% defined “Weak upregulation”. In the combined scores a logical AND combination was used that required all parameters to be fulfilled. For all three parameters 77 data sets from 21 ventilated COVID-19 patients were available. 17 ventilated patients were discharged from the ICU (survived) (69 samples) and 4 patients died on the ICU (8 samples). The number of right and false positive samples, the test sensitivity (Sens.), specificity (Spe.), negative (NPV) and positive predictive value (PPV) for predicting death are shown.

| **Stimulated blood:** |  |  |  |  |  |  |
| --- | --- | --- | --- | --- | --- | --- |
| **77 complete data sets of ventilated patients** | Right positive<br>(of 8 dead) | False Positive<br>(of 69 survived) | Sens. | Spe. | NPV | PPV |
| Weak upregulation of CD123 on Monos<br>+ weak upregulation of CD11b on Neutros<br>+ low Basophil count | 6 | 0 | 75% | 100% | 94% | 100% |
| Weak upregulation of CD11b on Neutros<br>+ low Basophil count | 6 | 1 | 75% | 99% | 94% | 93% |
| Weak upregulation of CD123 on Monos<br>+ weak upregulation of CD11b on Neutros | 7 | 3 | 88% | 96% | 97% | 83% |
| Weak upregulation of CD123 on Monos<br>+ low Baso count | 7 | 6 | 88% | 92% | 97% | 71% |
| Weak upregulation of CD11b on Neutros | 7 | 7 | 88% | 90% | 97% | 67% |
| Weak upregulation of CD123 on Monos | 8 | 15 | 100% | 80% | 100% | 52% |
| Low Baso count | 7 | 32 | 88% | 54% | 95% | 31% |

In accordance with published data^3,4^, male gender was more frequent in ventilated than non-ventilated patients (80% versus 52%). Ventilated patients also had a highly significant longer period of virus replication (28 days versus 10 days) that was defined as the period form the day of first clinical symptoms to the day of the last positive virus RT-PCR. Comorbidities were similar in both groups, while bacterial superinfection, thromboembolic complications and treatment with antibiotics or convalescent plasma were more frequent in ventilated patients (Tab. 1). As described before^27^, ventilated patients displayed higher plasma concentrations for biomarkers indicating inflammation (CRP, IL-6), liver and muscle dysfunction (LDH, Bilirubin, CK) and intravascular coagulation (D-dimer). Ventilated “dead” patients showed higher leukocyte counts and higher biomarkers for inflammation and liver dysfunction than ventilated “survived” patients (Supplementary Tab. 1).

### Impaired T cell reactivity is a hallmark of COVID-19 patients and correlates with disease severity

To measure T cell reactivity we used fresh heparinized whole blood and polyclonally activated the T cells with immobilized anti-CD3 antibodies. T cell activation was quantified not only by cytokine release but also by downstream effects on various cell types including basophils, pDCs, monocytes and neutrophils. In preparation of the study we have established that T cell activation results in a strong activation of basophils measured by upregulation of CD203c and downregulation of the IL-3-receptor beta-chain CD131 (used in this study). Basophils strongly express IL-3-receptors but only low levels of GM-CSF- and IL-5-receptors that also use CD131 for signal transduction^28^. Thus, among a variety of cytokines IL-3 is mainly responsible for downregulation of CD131 on basophils (Supplementary Fig. 1a). pDCs express IL-3- and GM-CSF-receptors; thus both cytokines contribute to downregulation of CD131 on pDCs (Supplementary Fig. 1b). CD14+ monocytes respond to T cell activation by a very pronounced upregulation of the IL-3-receptor alpha-chain CD123. Various cytokines, including IL-3, GM-CSF, IL-2, IL-15, IFN-gamma, IFN-alpha, TNF-alpha induce a strong upregulation of CD123 on monocytes (Supplementary Fig. 1c). Thus monocytes integrate a larger set of direct and indirect T cell-derived signals. Similarly, neutrophils respond to a specific set of cytokines with upregulation of CD11b (Supplementary Fig. 1d).

T cell activation with immobilized anti-CD3 in whole blood of healthy controls resulted in a pronounced downregulation of CD131 on basophils (to 13%) (Fig. 1a) as well as upregulation of CD123 on monocytes (to 1225%) (Fig. 1c) and CD11b on neutrophils (to 1232%) (Fig. 1e). In COVID-19 patients these changes were much weaker. Ventilated COVID-19 patients showed no significant downregulation of CD131 on pDCs, a weak downregulation of CD131 on basophils (to 77%), a weak upregulation of CD123 on monocytes (to 345%) and of CD11b on neutrophils (to 424%). Responses of non-ventilated patients were in-between the responses of ventilated patients and healthy controls. Persistence of an impaired T cell activation as defined by weak upregulation of CD123 und CD11b was significantly associated with prolonged viral replication in these patients (Supplementary Tab. 2). Absolute expression levels of surface markers on basophils, pDCs, monocytes and neutrophils and representative FACS dot plots of unstimulated and anti-CD3-stimulated whole blood cells are shown in Supplementary Figure 2a-d and Supplementary Figure 3. Downregulation of CD131 on basophils was almost completely dependent on IL-3 as shown with a blocking antibody against IL-3, while downregulation of CD131 on pDCs was only partially dependent on IL-3 (Supplementary Fig. 2a, b). Inhibition of IL-3 did not reduce the upregulation of CD123 on monocytes or CD11b on neutrophils (Supplementary Fig. 2c, d), most likely due to the high redundancy of T cell-derived signals with similar effects. Stratification of ventilated COVID-19 patients according to their clinical outcome revealed that downregulation of CD131 on pDCs or basophils was low in both groups (Fig. 1b). In contrast, upregulation of CD123 on monocytes and CD11b on neutrophils, that integrate a larger set of signals from activated T cells, were almost absent in COVID-19 patients with fatal outcome, but still detectable in ventilated patients who recovered and were later discharged from the ICU (Fig. 1d, f). In healthy males and non-ventilated male COVID-19 patients T cell activation resulted in a significantly stronger upregulation of CD123 on monocytes than in females, suggesting a stronger T cell - monocyte connection in males (Supplementary Fig. 4c). In ventilated COVID-19 patients the impairment of T cell-induced CD123 upregulation on monocytes was very pronounced in both genders (Supplementary Fig. 4c). Basophils, pDCs and neutrophils responded equally to T cell activation in males and females (Supplementary Fig. 4a, b, d).

**Fig. 1.**
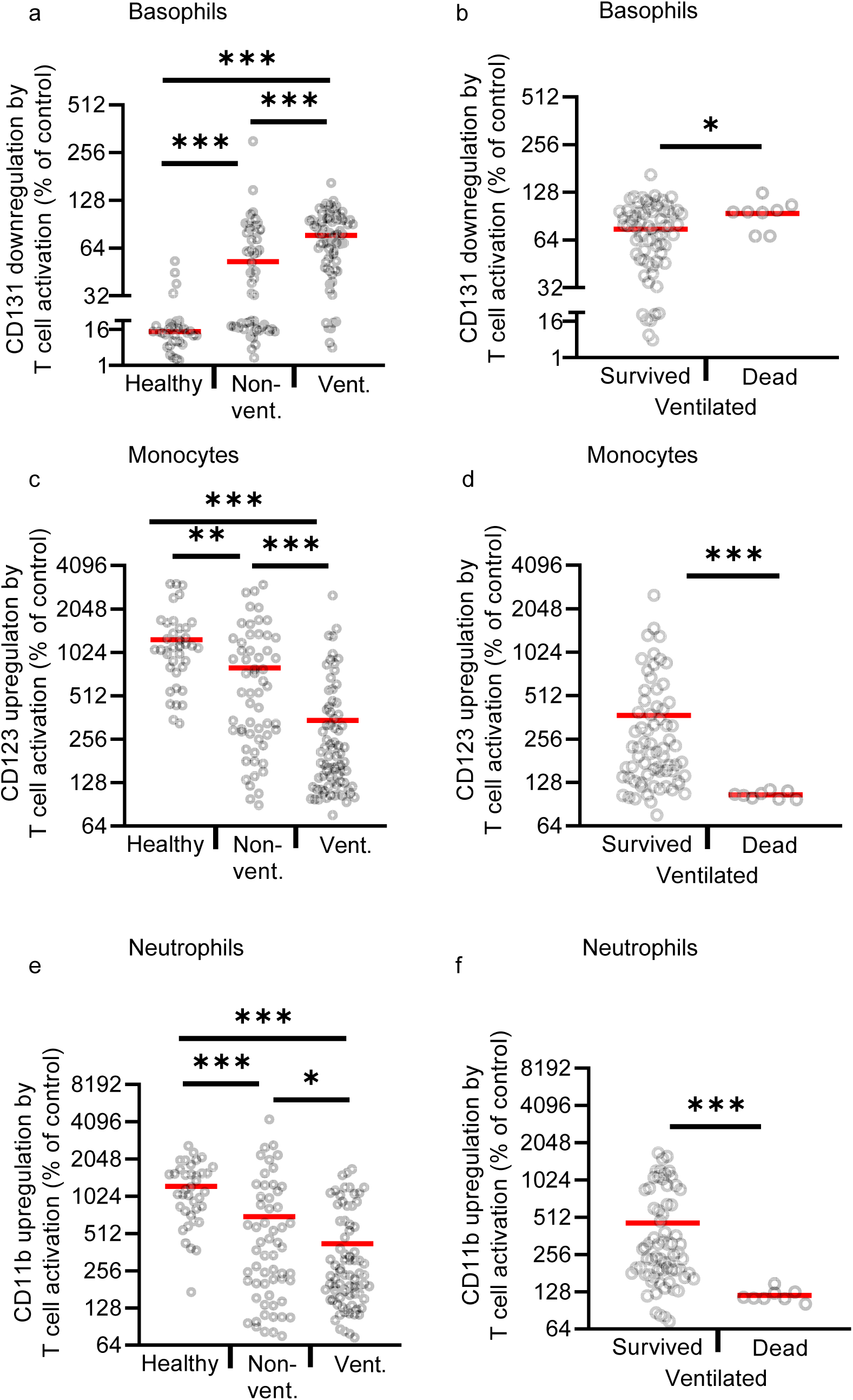
T cell reactivity in COVID-19 patients and healthy controls. **a**-**f** Whole blood samples from 38 healthy controls (Healthy; 38 samples), 33 non-ventilated (Non-vent.; 58 samples) and 21 mechanically ventilated (Vent.; 77 samples) COVID-19 patients were cultured with or without immobilized anti-CD3 for 24h. Ventilated patients were stratified into “survived” (17 patients, 69 samples) and “dead” (4 patients, 8 samples). Expression of indicated surface markers was quantified by flow cytometry on basophils (**a, b**), CD14+ monocytes (**c, d**) and neutrophils (**e, f**). Values depict the ratio of surface marker expression with anti-CD3 and surface marker expression without anti-CD3. Each sample is represented by one dot and the mean is marked in red. One-way ANOVA with Bonferroni multiple comparison test was used for **a, c**, and **e**. 2-tailed unpaired t-test with Welch’s correction was used for **b, d** and **f**. (^*^p<0.05,^**^p<0.01,^***^ p<0.001).

CD4+ and CD8+ T cell counts in whole blood samples were similar in ventilated patients, non-ventilated patients and healthy controls, arguing against the hypothesis that only differences in T cell numbers account for the differences observed in whole blood after T cell activation (Fig. 2a, b). Only in ventilated patients with fatal outcome CD4+ and CD8+ T cell counts were reduced.

**Fig. 2.**
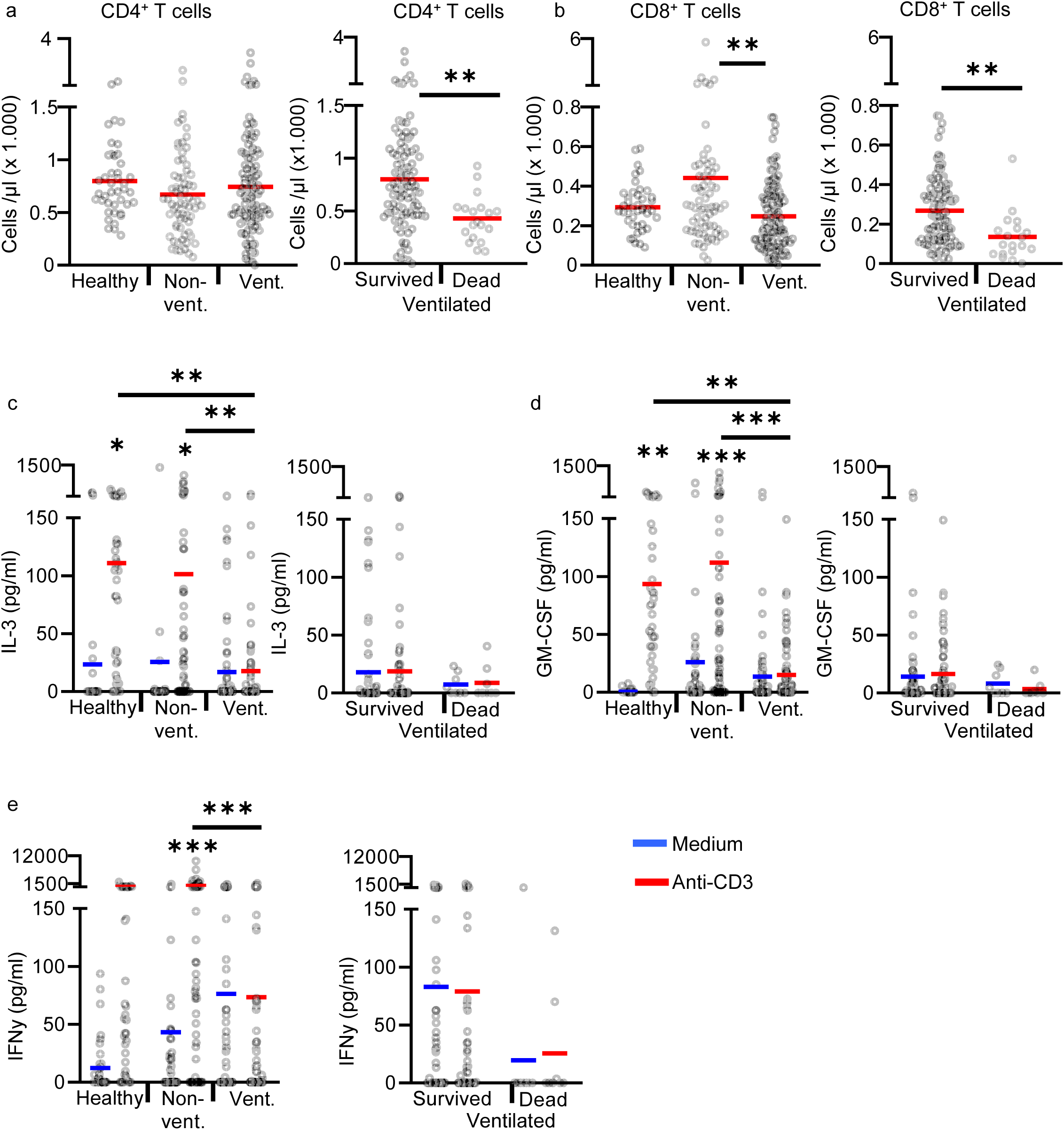
T cell counts and cytokine release in anti-CD3 activated whole blood samples. **a, b** Absolute CD4+ and CD8+ T cell counts in non-cultured fresh blood of 42 healthy controls (42 samples), 39 non-ventilated COVID-19 patients (68 samples) and 30 ventilated COVID-19 patients (120 samples). Ventilated patients were stratified into “survived” (23 patients, 101 samples) and “dead” (7 patients, 19 samples). Each sample is represented by one dot and the mean is marked in red. **c-e** Whole blood from 38 healthy controls (Healthy; 38 samples), 33 non-ventilated (Non-vent.; 58 samples) and 21 mechanically ventilated (Vent.; 77 samples) COVID-19 patients was cultured with or without immobilized anti-CD3 for 24h. Ventilated patients were stratified into “survived” (17 patients, 69 samples) and “dead” (4 patients, 8 samples). Concentrations of IL-3 (**c**), GM-CSF (**d**) and IFNγ (**e**) were measured in the culture supernatant by ELISA. Each sample is represented by one dot and the mean is marked in blue (Medium) or red (anti-CD3). One-way ANOVA with Bonferroni multiple comparison test was used. 2-tailed unpaired t-test was used for analysis of “Survived” and “Dead”. (^*^ p<0.05, ^**^ p<0.01, ^***^ p<0.001).

We also measured selected T cell-derived cytokines by ELISA in the supernatant of T cell activated whole blood (Fig. 2c-e). We found no significant anti-CD3-induced release of IL-3, GM-CSF and IFN-gamma in ventilated COVID-19 patients, but a considerable release in non-ventilated patients and healthy controls. These data are largely consistent with the above described downstream effects of T cell activation on basophils, pDCs, monocytes and neutrophils. However, the cytokine levels are highly variable and show no differences between non-ventilated COVID-19 patients and healthy controls. Thus, downstream cellular effects of T cell activation are more sensitive than measurement of single cytokines by ELISA.

### Whole blood is superior to PBMCs to quantify T cell anergy in COVID-19 patients

We also used PBMCs to polyclonally activate the T cells with soluble anti-CD3 for 24h. Downstream effects of T cell activation on pDCs and neutrophils could not be analyzed in PBMC samples, because neutrophils are absent and pDC numbers were too low. Basophils and monocytes responded only weakly to T cell derived signals without significant differences between the study groups (Supplementary Fig. 5a, b). Classical read-outs of T cell activation, like upregulation of CD25 on T cells (Supplementary Fig. 5c, d) and release of T cell-derived cytokines (Supplementary Fig. 5e) worked well and showed impaired T cell activation in ventilated COVID-19 patients with fatal outcome, but no significant differences between non-ventilated and ventilated patients. In order to analyze whether T cell anergy can be overcome with IL-2, we stimulated PBMC from patients with anti-CD3 alone or with a combination of anti-CD3 and IL-2 (Supplementary Fig. 5f). IL-2 significantly increased the release of GM-CSF and IFN-gamma, suggesting that IL-2 is able to partially overcome T cell anergy in COVID-19 patients.

### T cell anergy is reversible in critically ill COVID-19 patients

In several COVID-19 patients multiple blood samples were available over a prolonged period of time (Fig. 3). Four cases of ventilated patients that could finally be weaned from mechanical ventilation and two cases of patients that died on the ICU are shown. Patients with fatal outcome showed an almost complete T cell anergy in multiple samples over up to 14 days. In contrast, T cell anergy was less pronounced in patients with favorable outcomes. In general, their T cell reactivity appeared to improve already before weaning and was improved after successful weaning. This shows on an individual basis that T cell anergy is reversible and correlates to disease severity.

**Fig. 3.**
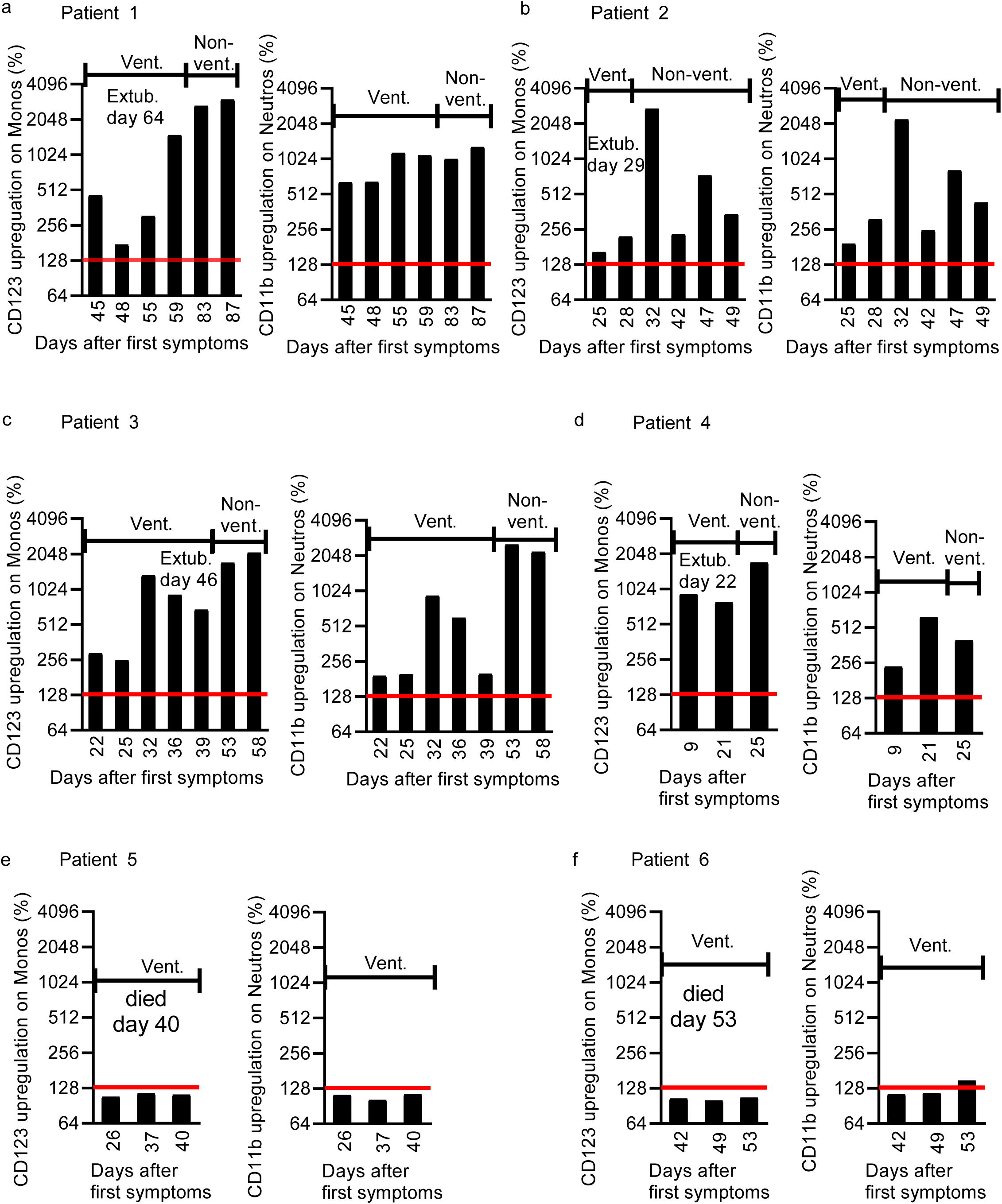
Examples of longitudinal analysis of ventilated COVID 19 patients. Upregulation of CD123 on CD14+ monocytes and CD11b on neutrophils in whole blood cultures with immobilized anti-CD3. Values depict the ratio of surface marker expression with anti-CD3 and surface marker expression without anti-CD3. 4 patients could be weaned from mechanical ventilation (patient 1-4) and 2 patients died on the ICU (patient 5-6). Periods of ventilation and non-ventilation are marked as “Vent.” and “Non-vent”. The cut-off of 130% upregulation that was used for the predictive score is shown as red line. Patient 1: 66-years old male with chronic kidney disease and diabetes. Patient 2: 68-years old woman with hypertension and atrial fibrillation. Patient 3: 64-years old woman with CLL stadium Binet B. Patient 4: 46-years old male with hypertension and obstructive sleep apnea. Patient 5: 68-years old male with hypertension and stable CLL. Patient 6: 68-years old male with hypertension, diabetes and prostatectomy due prostate cancer.

### Immunophenotyping of fresh blood samples in COVID-19 patients

We performed immunophenotyping of fresh blood samples of COVID-19 patients and healthy controls with a focus on innate cells like basophils, pDCs, monocytes and neutrophils (Fig. 4). The gating strategy is shown in Supplementary Fig. 6. Basophil counts were markedly reduced in ventilated COVID-19 patients that subsequently died on the ICU (Fig. 4a). Consistent with published data we also found decreased numbers of pDCs in ventilated COVID-19 patients (Fig. 4b).

**Fig. 4.**
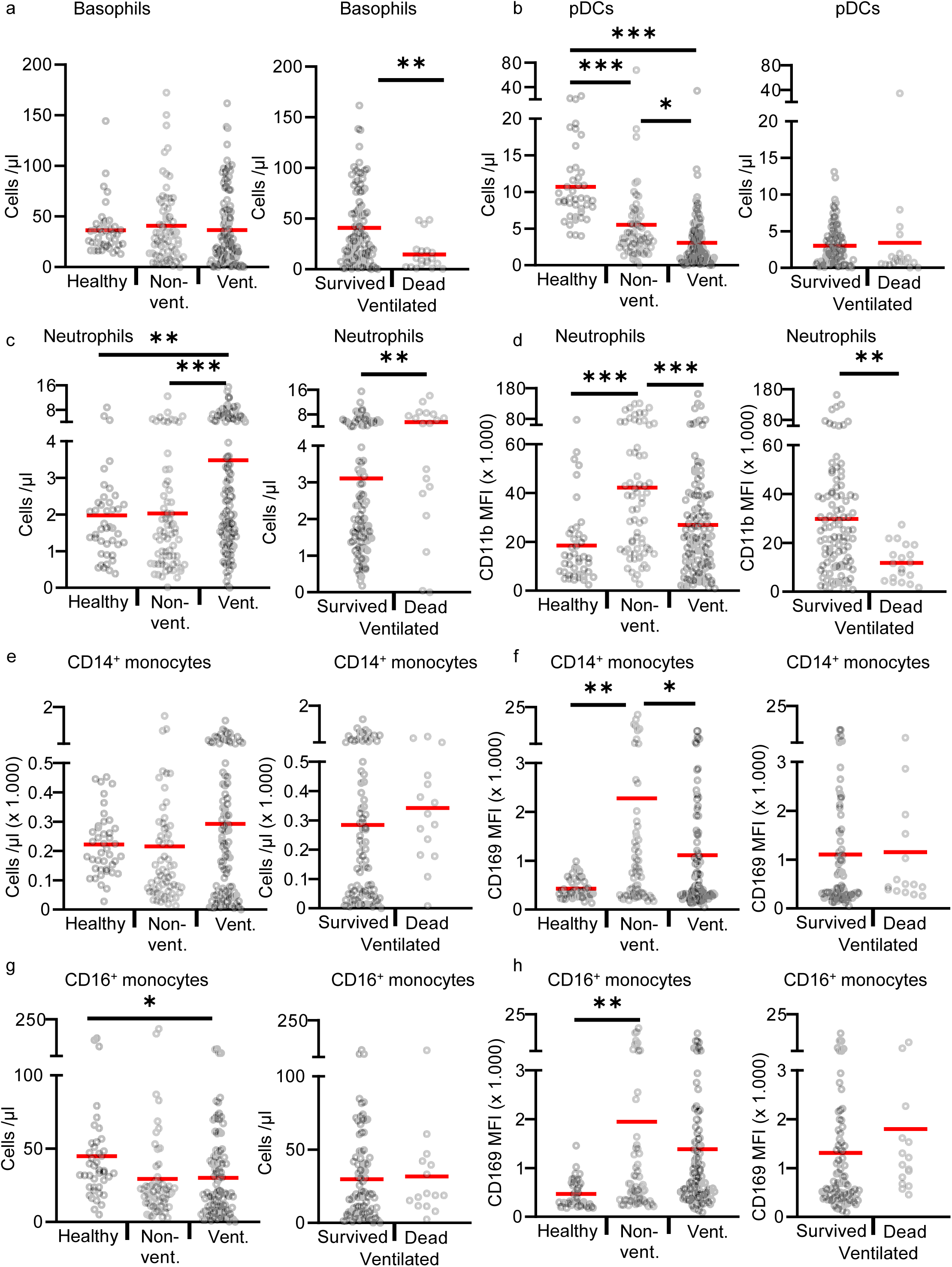
Immunophenotyping of fresh peripheral blood cells in COVID-19 patients and healthy controls. **a**-**d** 42 healthy controls (Healthy; 42 samples), 39 non-ventilated (Non-vent.; 68 samples) and 30 mechanically ventilated (Vent.; 120 samples) COVID-19 patients were analyzed. Ventilated patients were stratified into “survived” (23 patients, 101 samples) and “dead” (7 patients, 19 samples). **a, b** Absolute basophil and pDC counts. **c, d** Absolute neutrophils counts and their expression of CD11b given as mean fluorescence intensity (MFI). **e-h** 42 healthy controls (Healthy; 42 samples), 38 non-ventilated (Non-vent.; 62 samples) and 30 mechanically ventilated (Vent.; 102 samples) COVID-19 patients were analyzed. Ventilated patients were stratified into “survived” (23 patients, 87 samples) and “dead” (7 patients, 15 samples). The number of samples is lower than in a-d because the antibodies for this panel were temporarily not available by suppliers. **e, f** Absolute CD14++ CD16-monocyte counts and their surface expression of CD169. **g, h** Absolute CD16+ monocyte counts and their surface expression of CD169. Each sample is represented by one dot and the mean is marked in red. One-way ANOVA with Bonferroni multiple comparison test was used for analysis of “Healthy”, “Non-vent” and “Vent”. 2-tailed unpaired t-test was used for analysis of “Survived” and “Dead”. (^*^ p<0.05, ^**^ p<0.01, ^***^ p<0.001).

Total neutrophils counts were higher in ventilated COVID-19 patients and also higher in ventilated patients with a fatal outcome (Fig. 4c). Expression of CD11b on neutrophils showed the same pattern as CD169 on monocyte. CD11b was significantly higher in non-ventilated COVID-19 patients and returned almost to baseline in ventilated and deceased patients (Fig. 4d).

CD16+ monocytes but not the classical CD14+ monocytes were decreased in non-ventilated and ventilated patients (Fig. 4e, g). Both monocyte populations showed significantly increased expression of CD169 (Siglec-1) in non-ventilated patients compared to healthy controls (Fig. 4f, h). CD169 is one of the most prominent type I IFN-regulated genes on human monocytes^29^. In ventilated COVID-19 patients expression of CD169 returned almost to baseline, suggesting an impaired type I interferon production in critically ill COVID-19 patients. Reduced type I interferon responses were recently discovered by transcriptional profiling in critically ill COVID-19 patients^30^.

### Design and assessment of a predictive score

The immunological characteristics of COVID-19 patients lead us to develop predictive scores for survival of ventilated patients. We have incorporated three parameters that discriminated best between various groups of COVID-19 patients and calculated predictive values for death of ventilated patients (Tab. 2). The first two parameters are CD123-upregulation on monocytes and CD11b-upregulation on neutrophils induced by anti-CD3 in whole blood with a cut-off of 130%. Both parameters reflect the degree of T cell anergy; however they are not interchangeable, as monocytes and neutrophils sense different sets of T cell derived signals. The third parameter is the basophil count in the peripheral blood with a cut-off of 25 / µl. All three parameters should be easy to establish in different labs, because counting of basophils is routine and no standardized absolute expression levels are required for upregulation of CD123 and CD11b as only relative changes are calculated. By combination of all three parameters a positive predictive value of 100% and a negative predictive value of 94% were reached for fatal outcome of patients on mechanical ventilation.

## Discussion

We have shown that low T cell reactivity is a hallmark of COVID-19 patients and strongly correlates with disease severity. T cell reactivity was significantly lower in ventilated than in non-ventilated patients, and again significantly lower than in healthy controls.

Measurement of T cell activation by downstream effects on responder cells was much more sensitive and suitable to detect impairment of T cell activation and differences in patients than classical measurements of T cell-derived cytokines or T cell activation markers. The responder cells integrate a large set of signals from activated T cells in a cell type-specific manner. In addition, T cell activation assays with whole blood were superior to assays with purified PBMCs, because reactivity of basophils and monocytes was severely impaired after isolation of PBMCs. Whole blood has the advantage of little cell manipulation, a more natural environment within autologous plasma and the presence of all leukocyte subsets including neutrophils. With PBMCs and classical methods to study T cell activation conflicting data have been published showing unaltered, increased or decreased T cell responses in critically ill COVID-19 patients^20,21,23-26^.

Our data indicate that low T cell reactivity rather than anergy of downstream responder cells accounts for the low response of basophils, pDCs, monocytes and neutrophils, because relevant T cell-derived cytokines like IL-3, GM-CSF and IFN-gamma were reduced in a similar manner and neutralization of IL-3 markedly blocked the downstream effects on basophils.

The severe T cell anergy in critically ill COVID-19 patients does not seem to be a mere consequence of being critically ill, because T cell anergy was already seen in non-ventilated COVID-19 patients on a normal ward. Induction of T cell anergy could be a strategy of the virus to interfere with specific host immune responses and may explain the prolonged viral replication seen in the group of ventilated patients. Prolonged viral replication in turn may result in more tissue destruction, more activation of innate immune cells by viral components and more inflammation. The pronounced T cell anergy in critically ill patients with hyperinflammation strongly argues against a major direct role of T cells for hyperinflammation. Immunosuppressive approaches targeting T cells are unlikely to improve hyperinflammation, while strategies to overcome the pronounced T cell anergy in critically ill COVID-19 patients could be a promising therapeutic approach to improve virus clearance without aggravating the hyperinflammation.

Immunophenotyping revealed low counts of basophils and pDCs and a low expression of CD11b on neutrophils in severely ill COVID-19 patients. It is known that IL-3 is the major cytokine to increase basophil counts^28^. IL-3 and/or GM-CSF are also important survival factors for basophils and pDCs^31,32^. Thus the lower counts of pDCs and basophils in critically ill patients may reflect not only their increased recruitment into tissues but also reduced survival and production of cells due to diminished secretion of IL-3 and GM-CSF by anergic T cells. Also the lower expression of CD11b on neutrophils in ventilated patients may be explained by the more severe T cell anergy in ventilated patients.

Gender specific analysis showed that the T cell-induced upregulation of CD123 on monocytes was significantly higher in healthy males and non-ventilated male patients than in the same groups of females. As IL-3 contributes to activation of monocytes^33^, the stronger T cell – monocyte link in males may explain the recently described higher plasma levels of IL-8 and IL-18 in male COVID-19 patients^34^.

We have developed a score based on downstream T cell effects on monocytes and neutrophils, and absolute basophil counts to predict fatal outcome in ventilated COVID-19 patients. We would like to point out, that the score must be confirmed in additional cohorts of COVID-19 patients. The scores may be clinically useful to identify patients that may benefit from strategies to overcome T cell anergy.

Collectively our data suggest that approaches to overcome T cell anergy combined with concepts to specifically suppress hyperinflammation induced by innate immune cells, particularly monocytes/macrophages could be a promising immunotherapy for critically ill COVID-19 patients.

## Materials and Methods

### Study subjects and sampling

A total of 55 adult patients diagnosed with COVID-19 after testing positive for SARS-CoV-2 RNA by qPCR and 42 adult healthy volunteers without any clinical signs of COVID-19 infection were enrolled from March to July 2020 at the University Hospital Regensburg. This study was approved by the Research Ethics Committee from the University Hospital Regensburg (Study and Approval Number: 20-1785-101). Informed consent was obtained from all enrolled patients (or their legal representatives) and healthy volunteers.

The cohort of COVID-19 patients was stratified into non-ventilated (n=25) and mechanically ventilated (n=30) patients. Ventilated patients were further sub-grouped according to their outcome (discharged from the ICU = “survived”, or death on the ICU = “dead”) (Tab. 1). In most patients several consecutive blood samples were available resulting in a total of 188 samples. 14 patients were first sampled on mechanical ventilation and also after discharge from the ICU. Samples of these patients were assigned into the subgroup “ventilated” or “non-ventilated” according to the ventilation status at the time of sampling (Supplementary Tab. 1). None of the patients received prednisolone equivalents ≥ 40 mg/day for treatment of COVID-19.

Fresh whole blood samples for immediate immunophenotyping by flow cytometry with at least one antibody panel were available from all participants. Sufficient amounts of fresh blood to also perform whole blood stimulation were available from 38 healthy controls (38 samples), 33 non-ventilated COVID-19 patients (58 samples) and 21 mechanically ventilated COVID-19 patients (77 samples). For subgroup analysis the mechanically ventilated patients were again stratified according to their outcome into the groups “survived” (17 patients, 69 samples) and “dead” (4 patients, 8 samples). Peripheral blood mononuclear cells (PBMCs) were prepared from 25 non-ventilated patients (36 samples) and 16 ventilated COVID-19 patients (42 samples). For subgroup analysis the mechanically ventilated patients were again stratified according to their outcome into the groups “survived” (n=14, 39 samples) and “dead” (n=2, 3 samples). Exact numbers of patients and samples for each readout are provided in the figure legends, as for some read-outs not enough material was available.

### Cell culture media, cytokines and stimulants

For PBMC cultures we used RPMI 1640 medium (Gibco, Karlsruhe, Germany) containing 1% penicillin/streptomycin (Gibco, Karlsruhe, Germany) and 1% L-glutamine (Gibco, Karlsruhe, Germany). For whole blood stimulations we used pure RPMI 1640 medium (Gibco, Karlsruhe, Germany). Anti-CD3 (clone: OKT3) was obtained from eBiosciences (San Diego, CA) and used at 5 µg/ml. The blocking anti-IL-3 antibody (clone P8C11) was generated in our lab and used at a concentration of 10 µg/ml. IL-3 was obtained from BioLegend (San Diego, USA) and used at 20 ng/ml. GM-CSF, IL-2, IL-4, IL-5, IL-6, IL-15, IFN-γ, IFN-α, TNF-α were obtained from Peprotech (Cranbury, USA) and used at 20 ng/ml.

### Whole blood stimulation

For T cell activation assays tubes (BD Bioscience) were pre-coated with 300 µl anti-CD3 antibodies (5 µg/ml in PBS) at 37°C for 4 hours and washed twice with PBS. 100 µl of heparinized blood were diluted with 200 µl RPMI 1640 medium and added to the precoated or to uncoated tubes and incubated at 37°C in 5% CO_2_ for 24 hours. Where indicated a blocking anti-IL-3 antibody (10 µg/ml) was added to the samples. The supernatants were recovered and released cytokines analyzed by ELISA. Cells were analyzed by multi-parametric flow cytometry. For cytokine stimulation 100 µl whole blood was diluted with 200 µl RPMI 1640 medium and added to uncoated tubes (BD Bioscience) together with 20 ng/ml of cytokines (IL-2, IL-3, IL-4, IL-5, IL-6, IL-15, GM-CSF, IFN-γ, IFN-α and TNF-α) at 37°C in 5% CO_2_ for 24 hours. Cells were analyzed by flow cytometry.

### Preparation and culture of PBMCs

Peripheral blood mononuclear cells (PBMCs) were prepared by Ficoll-Paque density gradient centrifugation from heparin-anticoagulated fresh blood samples. For cryopreservation, PBMCs were resuspended in fetal calf serum (FCS) with 10% dimethylsulfoxide (DMSO) at a concentration of 1-2 x 10^6^ cells / ml, frozen at −80°C for 2-3 days and then transferred into liquid nitrogen. The cells were thawed in a 37°C water bath and washed 3 times with medium. The viability was controlled by Trypan-blue staining and was 90-95%. PBMCs (500.000/well) were cultured in 300 µl medium / well with or without anti-CD3 (clone: OKT3) 5 µg/ml for 24 hours by 37°C and 5% CO2. Where indicated IL-2 (20 ng/ml) was added to the samples during the incubation. Thereafter supernatants were recovered and analyzed by ELISA. Cells were analyzed by flow cytometry.

### Flow cytometry

Anti-coagulated fresh blood samples (100 µl) or cells after whole blood stimulations were incubated with various panels of the following directly labeled monoclonal antibodies for 20 minutes by 4°C: anti-CD3 APC-Cy7 (clone: SK7, BioLegend), anti-CD4 V500 (clone RPA-T4, BD Bioscience), anti-CD8 APC-eFluor 780 (clone: RPA-T8, Invitrogen, Darmstadt, Germany) and anti-CD8 PE-Cy7 (clone: SK1, BioLegend), anti-CD11b PE-Cy7(clone: M1/70, BioLegend, San Diego, CA), anti-CD14 V500 (clone: MΦP9, BD Bioscience), anti-CD16 Pacific Blue (clone: 3G8, BioLegend), anti-CD19 Pacific Blue (clone: HIB19, BioLegend), anti-CD25 APC (clone: BC96, BioLegend), anti-CD116 FITC (clone: 4H1, BioLegend), anti-CD131 PE (clone: 1C1, eBioscience), anti-CD123 PE-Cy5 (BD Bioscience), anti-CD169 PE (clone: 7-239, Miltenyi Biotec, Bergisch Gladbach, Germany), CD304 APC (clone: 12C2, BioLegend), HLA-DR II APC and FITC (clone: G-46-6, BD Bioscience). Subsequently, samples were treated with FACS Lysing Solution (BD Bioscience) for 10 min, washed with 0,9% NaCl, centrifuged, resuspended in 0,9% NaCl together with FACS counting beads (Invitrogen, Darmstadt, Germany) and acquired with a BD FACSCanto II flow cytometer and analyzed with FACSDIVA 8 software (BD Biosciences). For analysis of cultured PBMCs no erythrocyte lysis was performed. The gating strategy for key cell populations is shown in Supplementary Fig. 6.

### Enzyme Linked Immuno Sorbent Assay (ELISA)

Concentrations of GM-CSF and IFN-g in cell culture supernatants were measured by ELISA, according to manufacturer’s protocol (DuoSet Elisa, R&D Systems, Abington, UK). For measurement of IL-3, ELISA plates (NUNC Maxisorb, Thermofisher Scientific, Waltham, USA) were coated with a capture anti-IL-3 antibody (Clone P8C11; 5 µg/ml in PBS) in 100 µl / well at room temperature (RT) overnight. Plates were washed 3 times with PBS / 0.05 % Tween-20, blocked with PBS containing 1% bovine serum albumin (BSA) at RT for 1 hour and washed again with PBS. Samples where preincubated with 100 µg/ml mouse IgG1, kappa isotype control antibody (MOPC21, BioXCell, Lebanon, USA) for 1 hour by RT and added to the plates for 2 hours by RT (100 µl / well, diluted in PBS / 1% BSA). Recombinant human IL-3 (BioLegend) was diluted from 7.8 – 500 pg/ml in PBS / 1% BSA and served as standard. After washing plates were incubated with 400 ng/ml HRP-labelled detection anti-IL-3 antibody (Clone 13) (100 µl/well) for 1.5 hours at RT and color reaction was performed with TMB Substrate Solution (BioLegend) according to manufacturer’s protocol.

### Predictive score for death in ventilated COVID-19 patients

Three parameters were used individually or in combination to predict fatal outcome in ventilated COVID-19 patients. Absolute basophil counts in fresh peripheral blood. Basophil counts < 25 / µl defined “Low Baso count”. Upregulation of CD123 on CD14+ monocytes and CD11b on neutrophils was calculated as the ratio of surface marker expression with anti-CD3 and surface marker expression without anti-CD3. Upregulation of < 130% defined “Weak upregulation”. In the combined scores a logical AND combination was used that required all parameters to be fulfilled. For all three parameters 77 data sets from 21 ventilated COVID-19 patients were available. 17 ventilated patients were discharged from the ICU (survived) (69 samples) and 4 patients died on the ICU (8 samples). The number of right and false positive samples, the test sensitivity (Sens.), specificity (Spe.) was calculated. The negative (NPV) and positive predictive value (PPV) for predicting death were calculated using a prevalence of death in ventilated COVID-19 patients of 19 % (4 dead patients / 21 ventilated patients).

### Statistical analysis

Statistical analyses were performed using the GraphPad Prism 8 Software. Statistical differences between more than 2 different cell stimulations or patient cohorts were calculated using one-way

ANOVA with Bonferroni multiple comparison as indicated in the figures. For two group comparisons 2-tailed unpaired t-test was used as indicated in the figures. P values less than 0.05 were considered significant and marked with one asterisk or with two asterisks if less than 0.01. P values less than 0.001 were marked with three asterisks.

## Data Availability

The datasets generated during and/or analyzed during the current study are available from the corresponding author on reasonable request.

## Acknowledgements

This work was supported by a grant from the Bavarian Ministry of Science and Arts.

## Author contributions

M.M. conceived and supervised the study. K. R., M.M. analyzed data. C.M., C.T., JN. S., F.W., S. B. performed all experiments. M.V.M. provided patient data, M. L., D. L., B. G., F. H., F.H. B. S. provided patient samples and clinical data. H.P. obtained ethical approval for the study, M. K. provided healthy control samples. E. O., R. B. provided laboratory values and organized sample distribution. T. N., C. B., A. G. organized biobanking of PBMCs. M.M., K. R. drafted the manuscript.

## Competing interests

The authors declare no competing interests.

## Additional information

Supplementary information is available for this paper.

---

**Supplementary Fig. 1.**
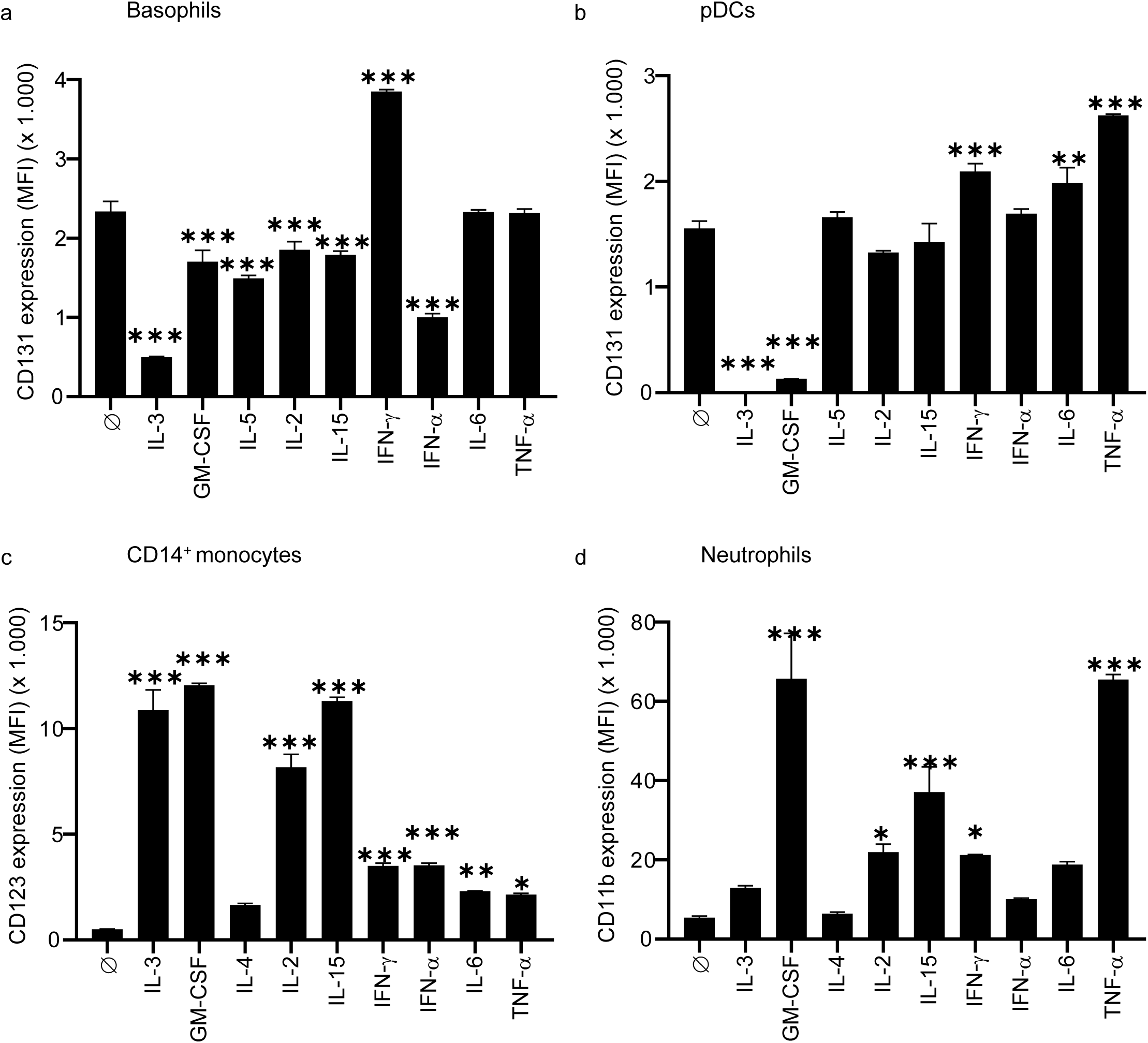
Cytokine induced changes of surface markers in a whole blood assay.. **a-d** Whole blood from a healthy donor was cultured in duplicates for 24 hours at 37°C with or without cytokines (IL-2, IL-3, IL-4, IL-5, IL-6, IL-15, GM-CSF, IFN-γ, IFN-α and TNF-α, 20 ng/ml each). Samples were analyzed by flow cytometry and absolute values of indicated cell surface markers are depicted as mean fluorescence intensity (MFI). **a** Expression of CD131 on basophils. **b** Expression of CD131 on pDCs. **c** Expression of CD123 on CD14+ monocytes. **d** Expression of CD11b on neutrophils. Data represent mean +/- standard deviation. Statistical differences between medium control and the various cytokines were calculated by one-way ANOVA with Bonferroni multiple comparison test (^*^ p<0.05, ^**^ p<0.01, ^***^ p<0.001).

**Supplementary Fig. 2.**
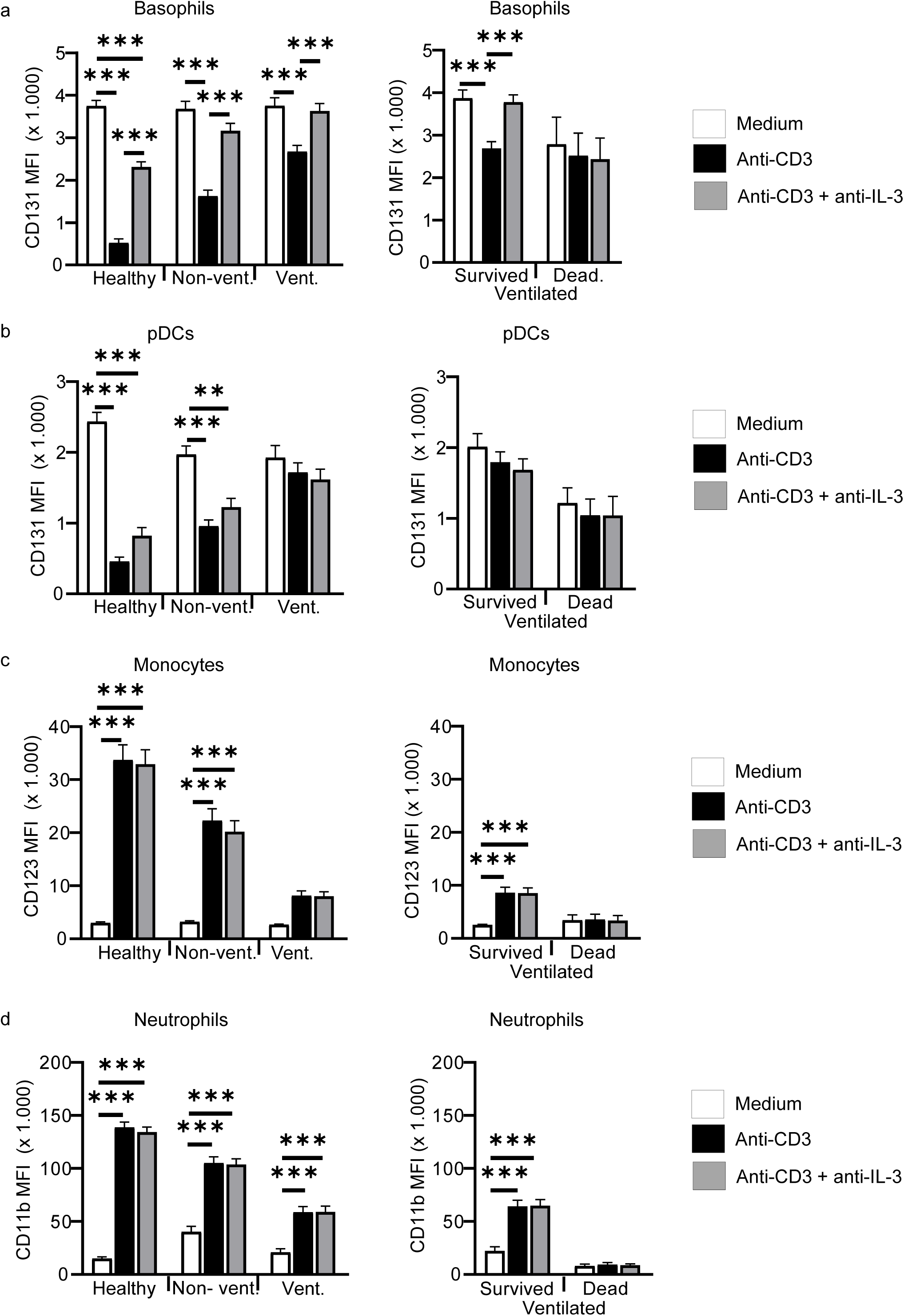
T cell reactivity in COVID-19 patients and healthy controls. **a-d** Whole blood from 38 healthy controls (Healthy; 38 samples), 33 non-ventilated (Non-vent.; 58 samples) and 21 mechanically ventilated (Vent.; 77 samples) COVID-19 patients was cultured without stimulation (medium), with immobilized anti-CD3, or with immobilized anti-CD3 plus anti-IL-3 (10 µg/ml) for 24h. Ventilated patients were stratified into “survived” (17 patients, 69 samples) and “dead” (4 patients, 8 samples). Expression of surface markers was quantified by flow cytometry and absolute expression values of indicated markers are shown as mean fluorescence intensity (MFI) on basophils (**a**), pDCs (**b**), CD14+ monocytes (**c**) and neutrophils (**d**). Bar graphs show mean +/- SEM (standard error of the mean). One-way ANOVA with Bonferroni multiple comparison test was used and statistical significance is only shown for differences between medium, anti-CD3 and anti-CD3+anti-IL-3 within each group of individuals. (^*^ p<0.05, ^**^ p<0.01, ^***^ p<0.001).

**Supplementary Fig. 3.**
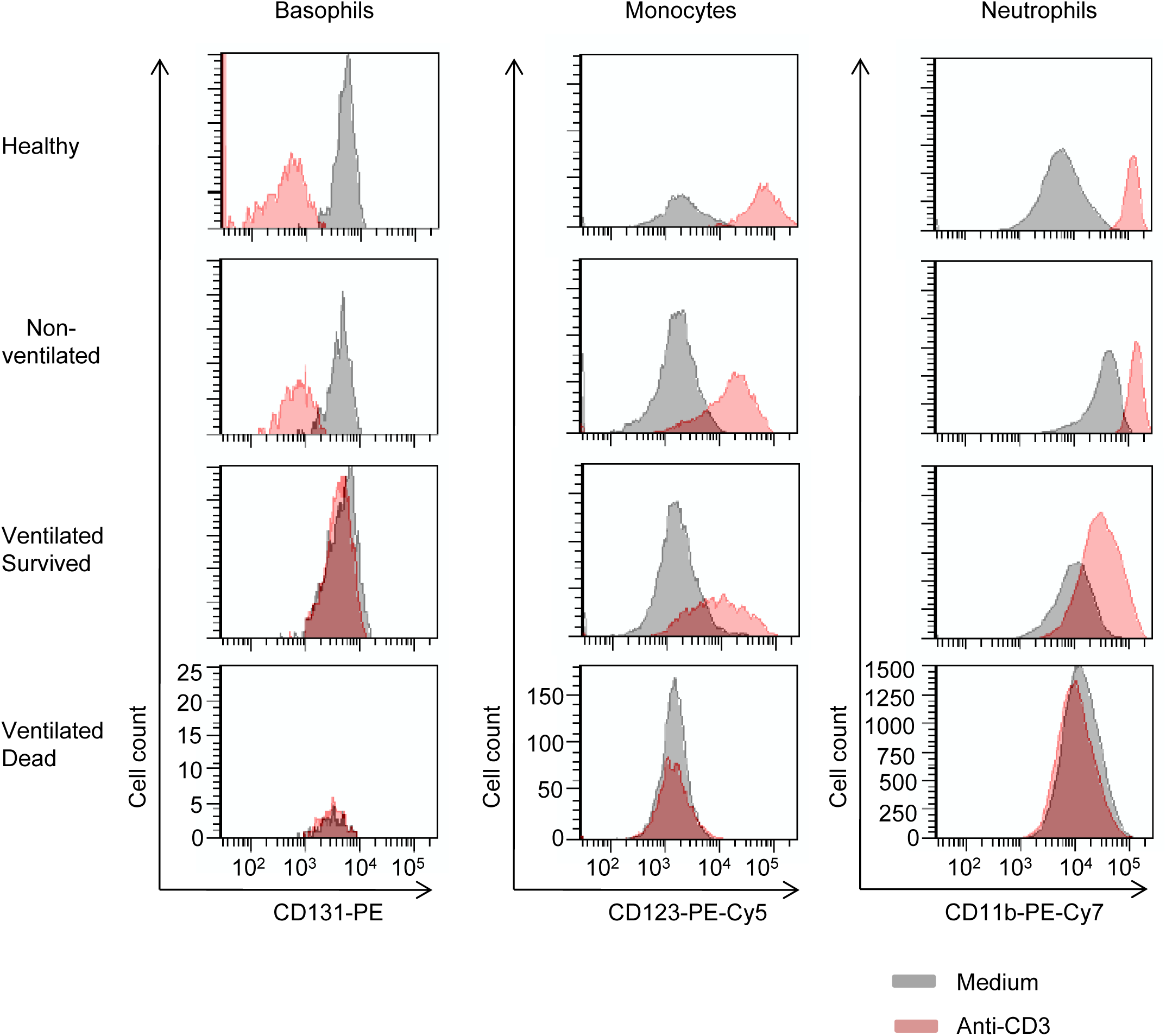
Representative FACS histogram plots for whole blood stimulation. Whole blood from various donors as indicated was cultured with (red) or without (grey) immobilized anti-CD3 for 24h. Expression of surface markers was quantified by flow cytometry and representative histogram plots are shown.

**Supplementary Fig. 4.**
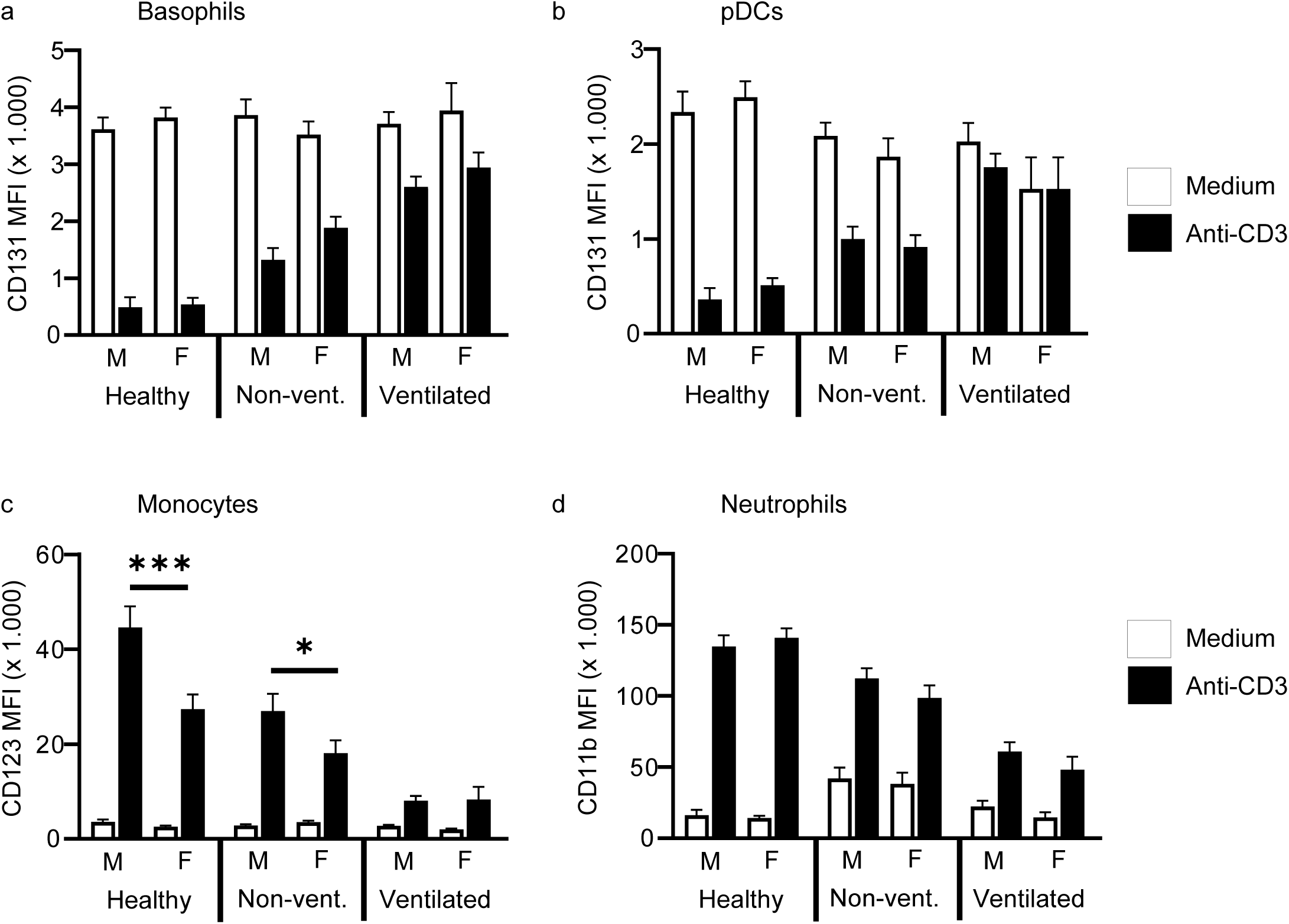
Gender specific analysis of T cell activation and immunophenotypes. **a-d** Whole blood from healthy controls (14 samples from 14 males and 24 samples from 24 females), non-ventilated COVID-19 patients (27 samples from 16 males and 31 samples from 17 females) and mechanically ventilated COVID-19 patients (62 samples from 16 males and 15 samples from 5 females) was cultured with or without immobilized anti-CD3 for 24h. Expression of surface markers was quantified by flow cytometry and absolute expression values of indicated markers are shown as mean fluorescence intensity (MFI) on basophils (**a**), pDCs (**b**), CD14+ monocytes (**c**) and neutrophils (**d**). Bar graphs show mean +/- SEM (standard error of the mean). One-way ANOVA with Bonferroni multiple comparison test was used and statistical significance is only shown for differences between males and femals within each group of individuals. (^*^ p<0.05, ^***^ p<0.001).

**Supplementary Fig. 5.**
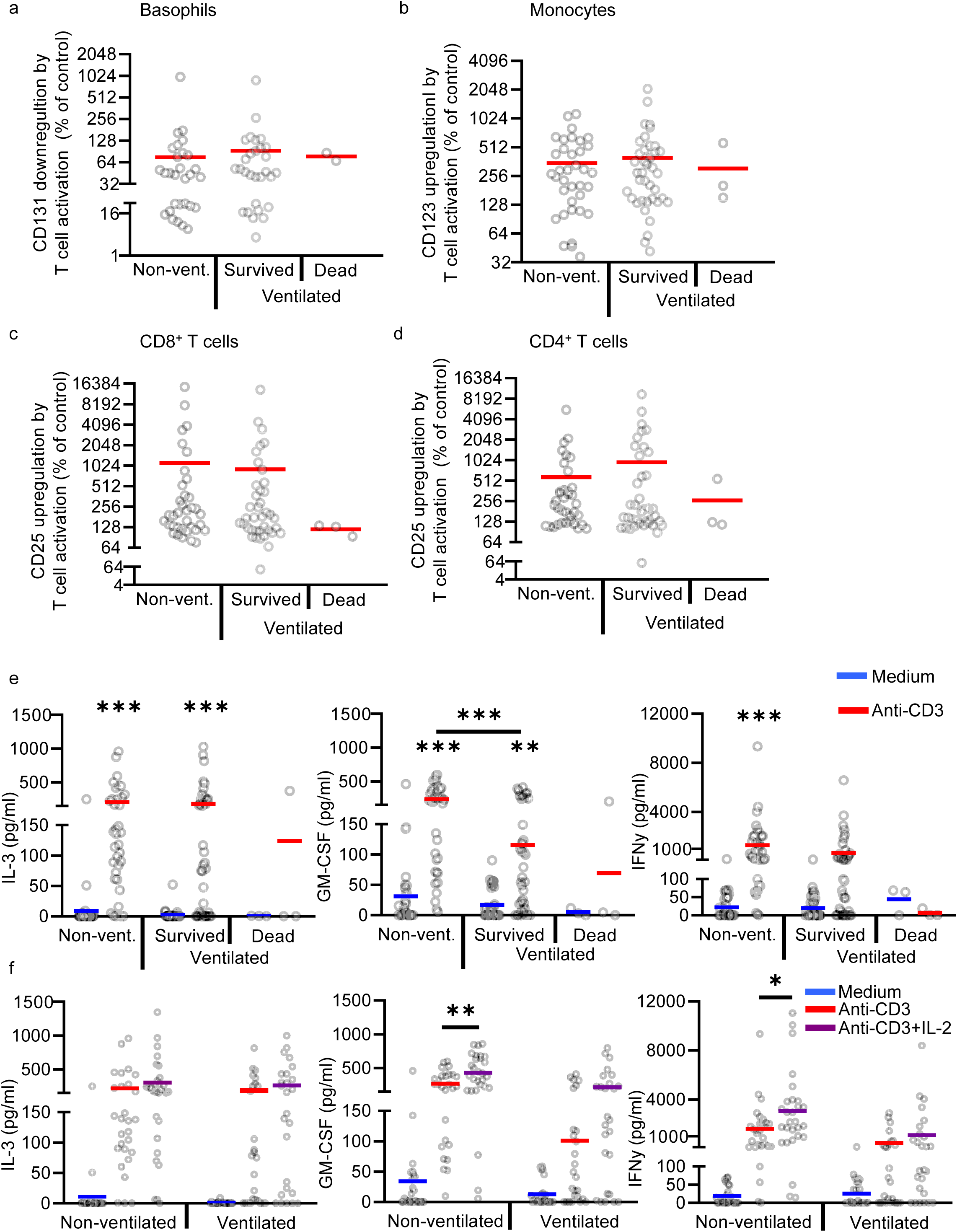
T cell reactivity analyzed with PBMC from COVID-19 patients. **a-e** PBMCs from 25 non-ventilated COVID-19 patients (Non-vent.; 36 samples), 14 mechanically ventilated COVID-19 patients that were discharged from the ICU (Ventilated Survived, 39 samples) and 2 mechanically ventilated COVID-19 patients that died on the ICU (Ventilated Dead, 3 samples) were cultured with or without anti-CD3 (5 µg/ml) for 24h. Analysis of basophils was not possible in all samples because basophil numbers were too low in some samples. Expression of indicated surface markers was quantified by flow cytometry on basophils (**a**), CD14+ monocytes (**b**), CD8+ T cells (**c**) and CD4+ T cells (**d**). Values depict the ratio of surface marker expression with anti-CD3 and surface marker expression without anti-CD3 in percent. Each sample is represented by one dot and the mean is marked in red. **e** Concentrations of IL-3, GM-CSF and IFNγ were measured in the culture supernatant by ELISA. Each sample is represented by one dot and the mean is marked in blue (Medium) or red (anti-CD3). One-way ANOVA with Bonferroni multiple comparison test was used. (^*^ p<0.05, ^**^ p<0.01, ^***^ p<0.001). **f** PBMCs from 22 non-ventilated COVID-19 patients (28 samples) and 12 ventilated COVID-19 patients (26 samples) were cultured with medium alone, with anti-CD3 or with anti-CD3 plus IL-2 (20 ng/ml) for 24h. Concentrations of IL-3, GM-CSF and IFNγ were measured in the culture supernatant by ELISA. Each sample is represented by one dot and the mean is marked in blue (Medium), red (anti-CD3) or purple (anti-CD3+IL-2). One-way ANOVA with Bonferroni multiple comparison test was used. (^*^ p<0.05, ^**^ p<0.01, ^***^ p<0.001).

**Supplementary Fig. 6.**
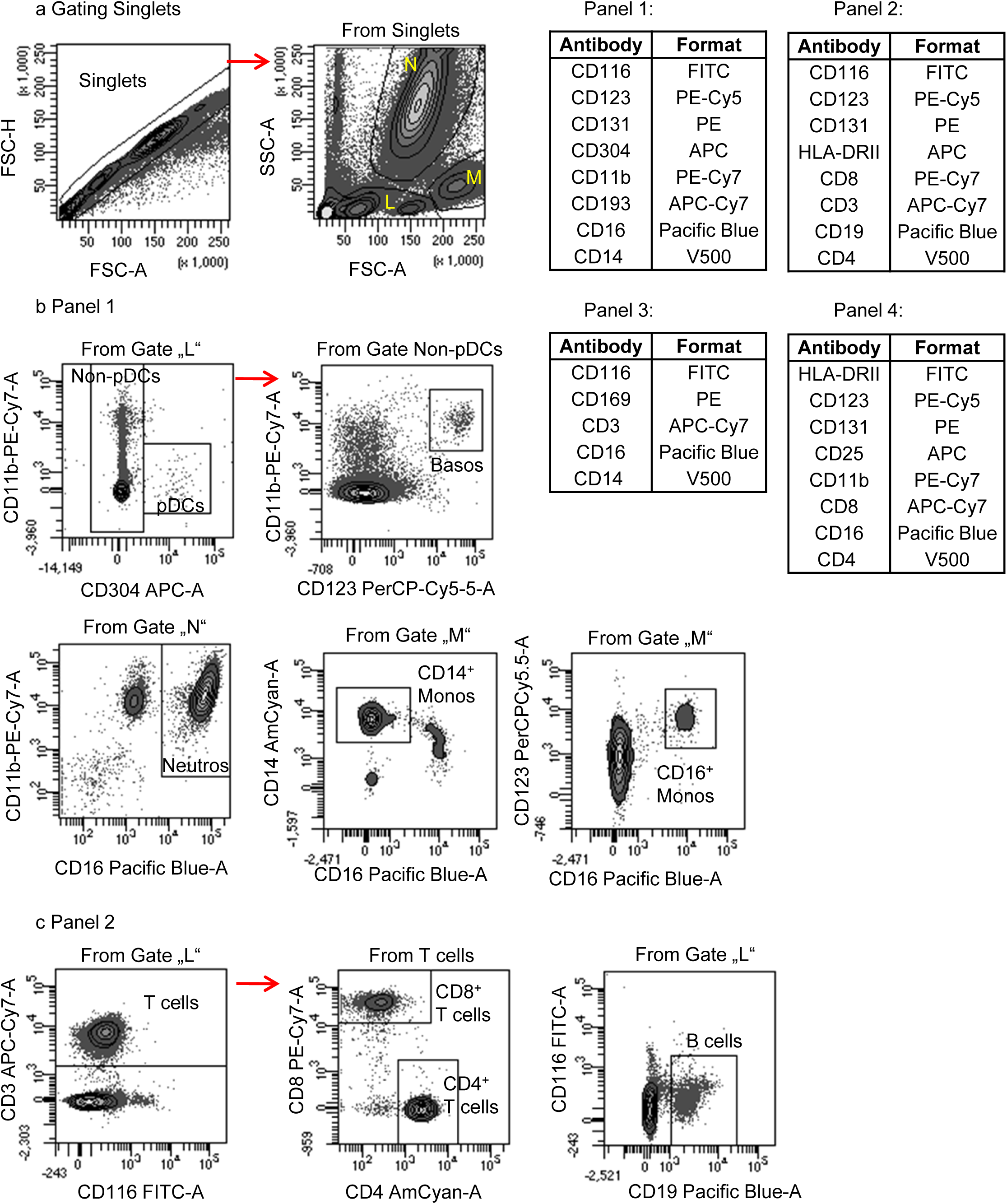
Gating strategy to detect key leukocyte subpopulations by flow cytometry. Gating strategy to identify pDCs, basophils, CD14+ monocytes, CD16+ monocytes (**a**), and T and B cells (**b**).

**Supplementary Tab. 1.** Clinical laboratory characteristics of COVID-19 patients and healthy controls. COVID-19 patients were subgrouped into non-ventilated and ventilated patients. Ventilated patients were further subgrouped into “Survived” and “Dead”. In most patients several consecutive blood samples were available. 14 patients were first sampled on ventilation and also after weaning from ventilation. 2-tailed unpaired t-test was used to calculate statistical differences between non-ventilated and ventilated patients as well as between “Survived” and “Dead” patients. 2-tailed unpaired t-test was used to calculate statistical differences (^*^ p<0.05, ^**^ p<0.01, ^***^ p<0.001).

|  | Healthy | Covid-19<br>all | Covid-19<br>non-ventilated | Covid-19<br>ventilated<br>all | Covid-19<br>ventilated<br>survived | Covid-19<br>ventilated<br>dead |
| --- | --- | --- | --- | --- | --- | --- |
| **Laboratory values:**<br>**(demographics)** |  |  |  |  |  |  |
| Number of samples (n) | 42 | 188 | 68 | 120 | 101 | 19 |
| Number of patients (n) | 42 | 55 | 39 # | 30 | 23 | 7 |
| **Laboratory values:**<br>**(mean)** |  |  |  |  |  |  |
| Procalcitonin (ng/ml) |  | 1.6 | 1.1 | 1.7 | 1.1 | 4.5 ** |
| CRP (mg/l) |  | 69.6 | 35.6 | 86.1 *** | 76.1 | 141,2 ** |
| IL-6 (pg/ml) |  | 85.0 | 23.9 | 94.3 | 71.1 | 216,4 *** |
| Ferritin (ng/ml) |  | 2721.9 | 1158.2 | 3018.5 | 1561.6 | 10950,4 *** |
| LDH |  | 340.2 | 273.9 | 370.6*** | 361.1 | 423,3 |
| ALAT (U/L) |  | 75.0 | 62.9 | 80.7 | 74.0 | 116,6 * |
| Bilirubin (mg/dl) |  | 2.2 | 0.9 | 2.8 * | 0.9 | 12,5 *** |
| CK (U/L) |  | 125.7 | 65.5 | 153.1 | 156.7 | 132,6 |
| D-Dimer (mg/L) |  | 8.2 | 4.0 | 8.6 * | 9.0 | 6,4 |
| Leucocytes (/nl) |  | 13.0 | 11,4 | 13.8 | 11.5 | 25,8 *** |

**Supplementary Tab. 2.** Correlation between persistence of SARS-CoV-2 replication and persistence of T cell anergy. Virus persistence was defined as the period from the day of first clinical symptoms to the last day of positive virus RT-PCR. Patients were stratified in two groups by virus persistence < 15 days (n=18) or ≥ 15 days (n=25). T cell reactivity was quantified by upregulation of CD123 on monocytes or CD11b on neutrophils as described in Fig. 1. T cell anergy was defined as an upregulation < 300% for both surface markers. Persistence of T cell anergy was defined as the period from the day of first clinical symptoms to the last day of T cell anergy. 2-tailed unpaired t-test was used to calculate statistical differences between the two groups of patients. (^*^ p<0.05, ^**^ p<0.01, ^***^ p<0.001).

|  | Virus persistence<br>in days<br>(mean +/- SEM) | Persistence of T cell anergy<br>in days (mean +/- SEM),<br>measured by CD123<br>upregulation on monocytes | Persistence of T cell anergy<br>in days (mean +/- SEM),<br>measured by CD11b<br>upregulation on neutrophils |
| --- | --- | --- | --- |
| Patients with virus<br>persistence < 15 days | 5.8 (± 1.1) | 5.3 (± 2.7) | 10.6 (± 3.3) |
| Patients with virus<br>persistence ≥ 15 days | 31.0 *** (± 3.1) | 24.0 ** (± 4.8) | 23.9 * (± 4.6) |

## Notes

### Competing Interest Statement

The authors have declared no competing interest.

### Author Declarations

Research Ethics Committee from the University Hospital Regensburg (Study and Approval Number: 20-1785-101)

